# Characterization of a high-resolution breath acetone meter for ketosis monitoring

**DOI:** 10.1101/2020.04.20.20072975

**Authors:** Donald J. Suntrup, Timothy V. Ratto, Matt Ratto, James P. McCarter

## Abstract

**Background:** The ketone bodies beta-hydroxybutyrate (BHB) and acetone are endogenous products of fatty acid metabolism. Although ketone levels can be monitored by measuring either blood BHB or breath acetone, determining the precise correlation between these two measurement methods has been challenging. The purpose of this study is to characterize the performance of a novel portable breath acetone meter (PBAM) developed by Readout, Inc., to compare single versus multiple daily ketone measurements, and to compare breath acetone (BrAce) and blood BHB measurements.

**Methods:** We conducted a 14-day prospective observational cohort study of 21 subjects attempting to follow either a low-carbohydrate/ketogenic or a standard diet. Subjects were asked to concurrently measure both blood BHB and BrAce five times per day and report the results using an online data entry system. We evaluated the utility of multiple daily measurements by calculating the coefficient of variation (CV) for each daily group of measurements. We calculated the correlation between coincident BrAce and blood BHB measurements using linear ordinary least squares regression analysis. We assessed the ability of the BrAce measurement to accurately predict blood BHB states using receiver operating characteristic (ROC) analysis. Finally, we calculated a daily ketone exposure (DKE) using the area under the curve (AUC) of a ketone concentration versus time graph and compared the DKE of BrAce and blood BHB using linear ordinary least squares regression.

**Results:** BrAce and blood BHB varied throughout the day by an average of 44% and 46%, respectively. The BrAce measurement accurately predicted whether blood BHB was greater than or less than the following thresholds: 0.3 mM (AUC = 0.898), 0.5 mM (AUC = 0.854), 1.0 mM (AUC = 0.887), and 1.5 mM (AUC = 0.935). Coincident BrAce and blood BHB measurements were moderately correlated with *R*^2^ = 0.57 (*P* < 0.0001), similar to literature reported values. However, daily ketone exposures, or areas under the curve, for BrAce and blood BHB were highly correlated with *R*^2^ = 0.83 (*P* < 0.0001).

**Conclusions:** The results validated the performance of the PBAM. The BrAce/BHB correlation was similar to literature values where BrAce was measured using highly accurate lab instruments. Additionally, BrAce measurements using the PBAM can be used to predict blood BHB states. The relatively high daily variability of ketone levels indicate that single blood or breath ketone measurements are often not sufficient to assess daily ketone exposure for most users. Finally, although single coincident blood and breath ketone measurements show only a moderate correlation, possibly due to the temporal lag between BrAce and blood BHB, daily ketone exposures for blood and breath are highly correlated.

## INTRODUCTION

Ketone bodies (“ketones”) are endogenous products of liver fatty acid metabolism. In humans, ketones rise from low basal levels during states of prolonged fasting or carbohydrate restriction when low insulin levels and increased free fatty acid (FFA) concentrations lead to an upregulation of ketogenesis. Ketones, along with FFAs, are released into the blood and enter cells serving as alternative energy substrates to glucose. Although all non-hepatic cells can metabolize ketones, they are particularly utilized by the heart and the brain. Ketones also have signaling properties including inhibition of pathways for inflammation (Youm et al., 2015) and oxidative stress (Shimazu et al., 2013).

The state in which the body derives the majority of its energy from ketone and FFA metabolism is called ketosis. Nutritional and fasting ketosis, where ketone levels are elevated but moderate, are physiologically distinct from diabetic ketoacidosis (DKA), an acute pathological condition characterized by runaway ketogenesis and metabolic acidosis. While DKA poses a risk for insulin-dependent diabetics, there is no risk associated with nutritional and fasting ketosis.

For decades, clinicians and researchers have deliberately induced the state of ketosis to treat a variety of pathological conditions. The 1920s saw the first clinical use of the ketogenic diet for the treatment of severe epilepsy (Paoli et al., 2013). More recently, nutritional ketosis has been used as a successful therapy for the management of type-2 diabetes (McKenzie et al., 2017; Hallberg et al., 2018), metabolic syndrome (Hyde et al., 2019) and obesity (Brehm et al., 2003; Shai et al., 2008). Evidence is emerging that the ketogenic diet may be effective either as a primary or adjunct therapy for polycystic ovary syndrome (Alwahab et al., 2018), cancer (Hopkins et al., 2018), migraine headaches (Di Lorenzo et al., 2019) and other neurological diseases (Taylor et al., 2019). Although the mechanisms behind these benefits are not well understood, researchers have begun exploring hypotheses (Elamin et al., 2017).

In all of these clinical applications, regularly monitoring ketone levels helps gauge patient adherence to prescribed dietary protocols. Furthermore, for a fixed dietary protocol, differences in basal metabolic rate, hepatic glycogen stores and other factors lead to differences in ketone production rates across individuals (Mitchell et al., 1995). Therefore, monitoring ketone levels helps individuals and their caregivers understand the personal propensity for ketone production, especially in response to dietary and lifestyle changes.

There are three primary methods for measuring ketones, each of which is sensitive to one of the three major ketone bodies: acetoacetate (AcAc), beta-hydroxybutyrate (BHB) and acetone. Figure 1 summarizes the production, metabolism and excretion of the three ketone bodies and how they are interrelated. During periods of carbohydrate restriction, acetyl-CoA derived from FFAs in the liver is diverted to produce AcAc. A portion of the AcAc is converted to BHB, with the specific ratio determined by the redox potential (i.e. NADH/NAD^+^) of the liver mitochondria and the available supply of the catalyzing enzyme D-beta-hydroxybutyrate dehydrogenase (3-HBDH) (Laffel, 1999).

**Figure 1.**
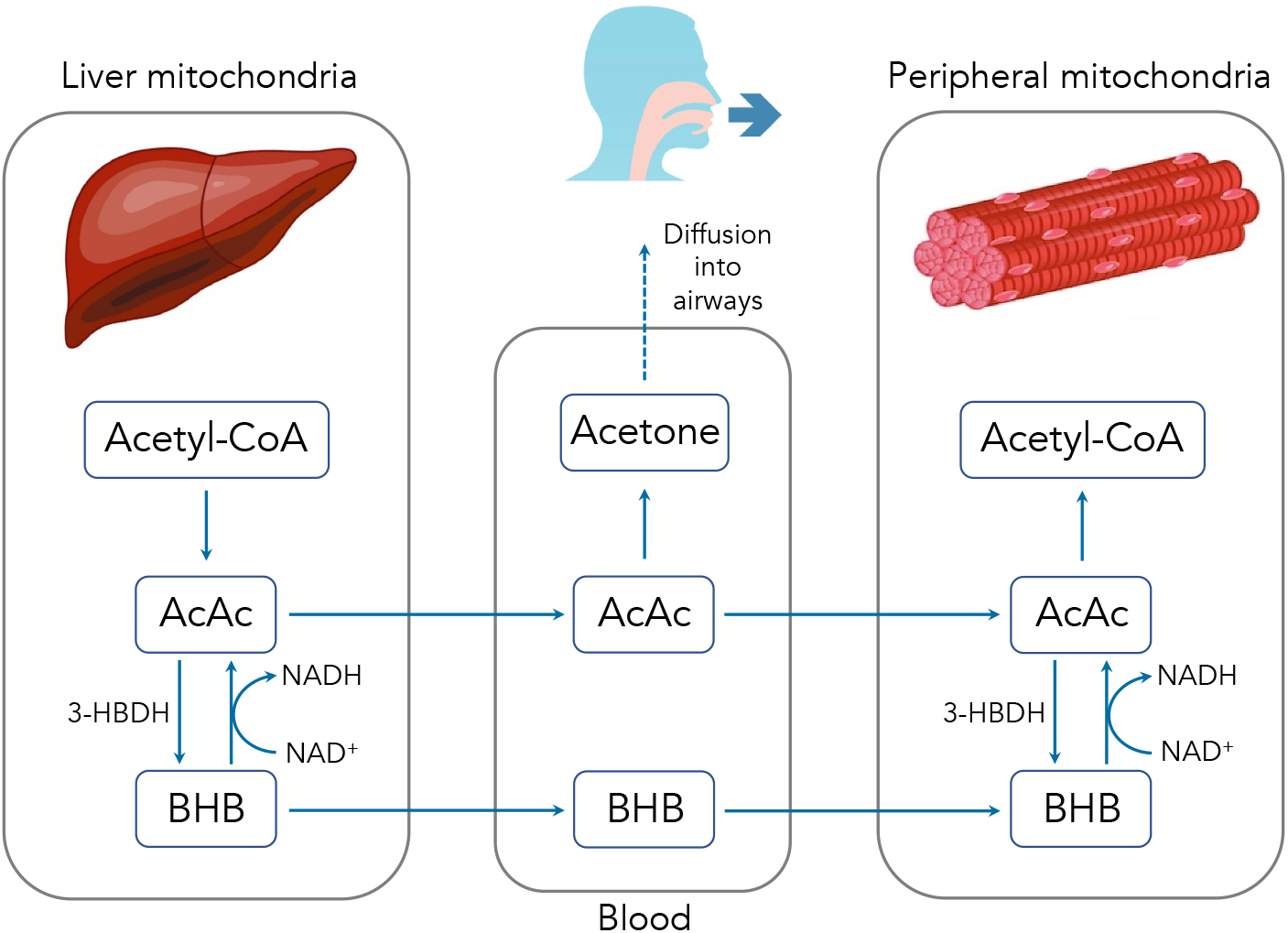
Summary of ketone body production, metabolism and excretion.

AcAc and BHB are released into the bloodstream where they can travel to peripheral cells and serve as metabolic substrates. Once inside the peripheral cell mitochondria, AcAc and BHB may interconvert through the action of 3-HBDH. Intracellular AcAc can then be converted into acetyl-CoA and used for metabolism. Excess AcAc is excreted in the urine where it can be measured using urinary test strips via the nitroprusside reaction. BHB is measured in the blood using a finger prick and test strip system. Circulating AcAc spontaneously degrades via decarboxylation into acetone. Owing to its small size and volatility, acetone diffuses freely into the airways and is exhaled in the breath. Because acetone is a direct derivative of AcAc, accurately determining the concentration of breath acetone (BrAce) provides a reliable indication of the depth of ketosis (Musa-Veloso et al., 2002).

Studies have demonstrated that BrAce becomes elevated during states of carbohydrate restriction, caloric restriction and fasting in diabetic and obese subjects (Freund, 1965; Tassopoulos et al., 1969). Because BrAce rises any time fatty acids are metabolized to meet energy demands, several studies have found a strong correlation between the specific BrAce concentration and the rate of body fat loss (Kundu, 1990; Kundu et al., 1993; Landini et al., 2007). Other studies have concurrently measured both BrAce and blood BHB and have found correlation coefficients (*R*^2^) ranging from 0.54−0.94 with a weighted mean of *R*^2^ = 0.64 (n = 506) (Anderson, 2015; Musa-Veloso et al., 2006; Tassopoulos et al., 1969; Rooth and Carlström, 1970; Musa-Veloso et al., 2002; Prabhakar et al., 2014; Qiao et al., 2014; Güntner et al., 2018). In the majority of these studies, BrAce was measured using laboratory methods including gas chromatography (GC-MS/FID) and selected-ion flow-tube mass spectrometry (SIFT-MS). Although these diagnostic tools are highly sensitive and selective, their size, cost and training requirements prevent their use as personal monitoring devices.

An alternative solution for portable BrAce monitoring is based on chemoresistive metal oxide semi-conductor (MOS) sensors. A well designed MOS sensor system can enable highly accurate BrAce monitoring in a handheld device. Because of the portability and noninvasive nature of such a device, simple, user-friendly high-frequency ketone monitoring becomes feasible. Until now, however, the accuracy of portable BrAce monitors has been limited by overly simplistic device design and the absence of an appropriate breath sampling method.

Readout Health has developed a high-resolution portable breath acetone meter (PBAM) for ketosis monitoring (Figure 2a). The PBAM contains a MOS sensor that is highly selective to acetone over the most common interfering analytes in breath (e.g. hydrogen, alcohols). Furthermore, the PBAM relies on a modified vital capacity maneuver using a pressure sensor and pump to selectively sample only the latter portion of the exhalation (i.e. alveolar gas) where the concentration of acetone is highest and most repeatable. Lastly, the PBAM sensor is housed inside a sealed flow cell, which eliminates the confounding effect of ambient air mixing with the breath sample during a measurement.

**Figure 2.**
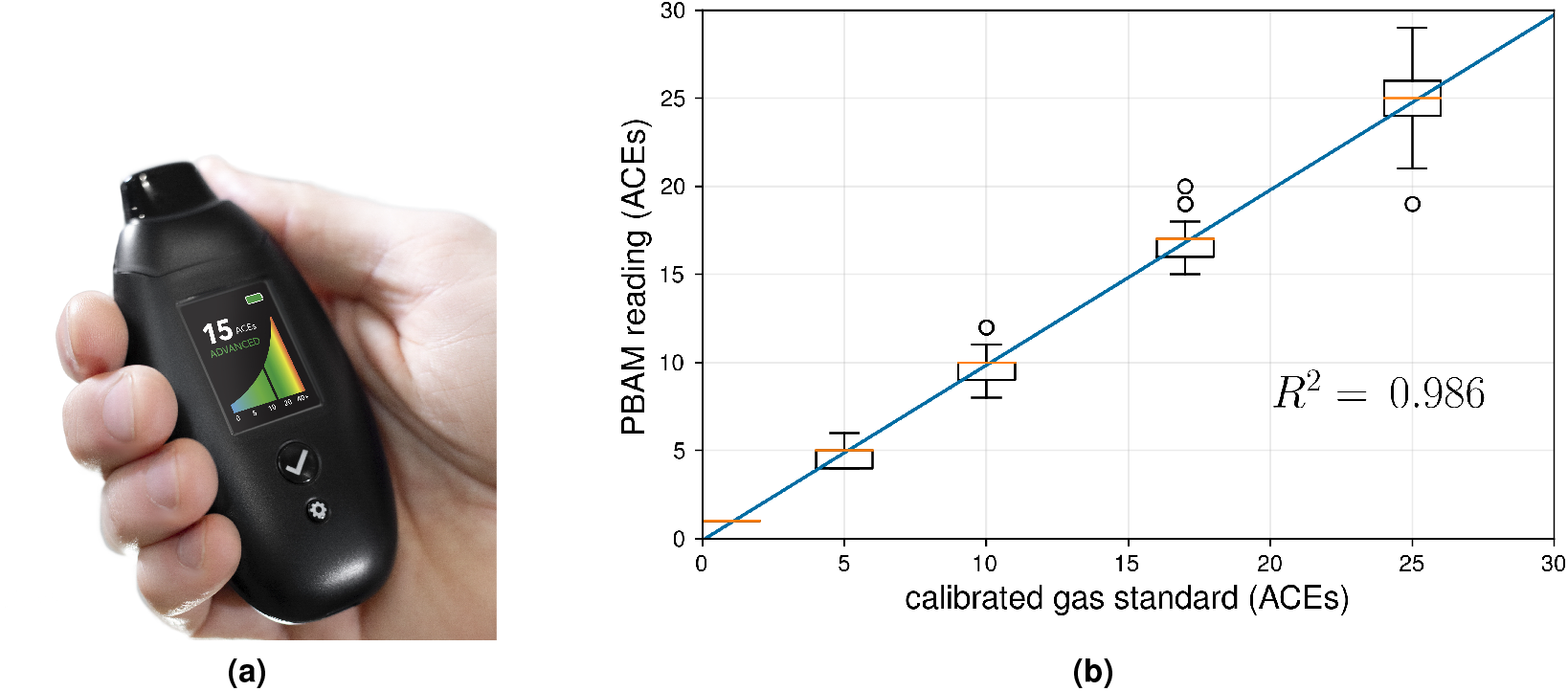
(a) The PBAM developed by Readout Health (b) Performance of three calibrated PBAM’s against a laboratory gas standard. The readings from the PBAM and the gas standard were linearly correlated with an *R*^2^ of 0.986. The orange line indicates the median and the box edges represent the 25^th^ quartile (*Q*_1_) and 75th quartile (*Q*_3_) for each gas concentration. The box width represents the interquartile range (IQR = *Q*_3_ *− Q*_1_).The upper and lower whiskers represent the last datum less than *Q*_3_ + 1.5 *\** IQR and the first datum greater than *Q*_1_ *−* 1.5 *\** IQR, respectively. Finally, the open circles represent data beyond *Q*_3_ + 1.5 *\** IQR and *Q*_1_ *−* 1.5 *\** IQR.

The PBAM reports BrAce results in units called ACEs, which are designed to translate parts per million (ppm) of BrAce into a blood BHB equivalent. As blood BHB and BrAce are enzymatically and non-enzymatically converted from AcAc, and BrAce is additionally dependent on blood-gas partitioning, the relationship between BrAce and BHB is nonlinear. Based on the literature and our own data (Musa-Veloso et al., 2002) the relationship can be described by a function of the form:

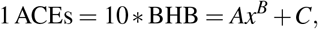

where *x* is BrAce in ppm and *A, B* and *C* are device specific coefficients.

Each PBAM is individually calibrated using a laboratory gas acetone standard at three different concentrations. After calibration, each PBAM is exposed to the lab acetone standards a second time and screened for accuracy. Figure 2b shows the performance of three calibrated PBAMs upon repeated exposure to the laboratory acetone standards. The results demonstrate high accuracy and repeatability after individual calibration.

The purpose of this clinical study is to characterize the PBAM and to compare BrAce measurements from the PBAM to blood BHB. In addition, we investigate the utility of a single daily ketone measurement compared to multiple daily measurements. Finally, we introduce the concept of daily ketone exposure (DKE) and compare DKE for BrAce and blood BHB.

## MATERIALS & METHODS

### Design and Subjects

We conducted a 14-day prospective observational cohort study of 21 subjects attempting to follow a low-carbohydrate/ketogenic or standard diet. Both male and female subjects over the age of 18 and from all ethnic groups were considered for both cohorts of the study. Subjects were recruited based on the diet they were following prior to the study. Therefore, none were asked to make dietary changes. Subjects were recruited with printed advertisements and word of mouth and were evaluated via email or phone. The duration of the recruitment period was 6 weeks.

Candidates with type 1 diabetes, insulin-dependent type 2 diabetes or a history of diabetic ketoacidosis (DKA) were excluded as well as those taking warfarin (blood thinners), sodium-glucose cotransporter-2 (SGLT2) inhibitors and disulfiram. One subject had type-2 diabetes but was not insulin dependent.

Subjects were asked to monitor their blood and breath ketones five times per day. The purpose of the two cohorts was to generate the broadest range of ketone values across the entire group. In addition to the standard diet (high-carbohydrate) subjects, several subjects in the ketogenic/low-carb diet arm failed to achieve elevated ketone levels and thus served as additional sources of low ketone data. Table 1 describes the gender and age distributions of the study subjects, and Figure 3 shows the study flow.

**Table 1.** Descriptive data of the 21 subjects in the ketogenic/low-carb and standard diet arms.

| Cohort | n | Gender |  | Age |  |
| --- | --- | --- | --- | --- | --- |
|  |  | Male | Female | Mean | SD |
| Ketogenic/Low-carb | 19 | 21% | 79% | 43.6 | 14.4 |
| Standard mixed | 2 | 0% | 100% | 43 | 21.2 |

**Figure 3.**
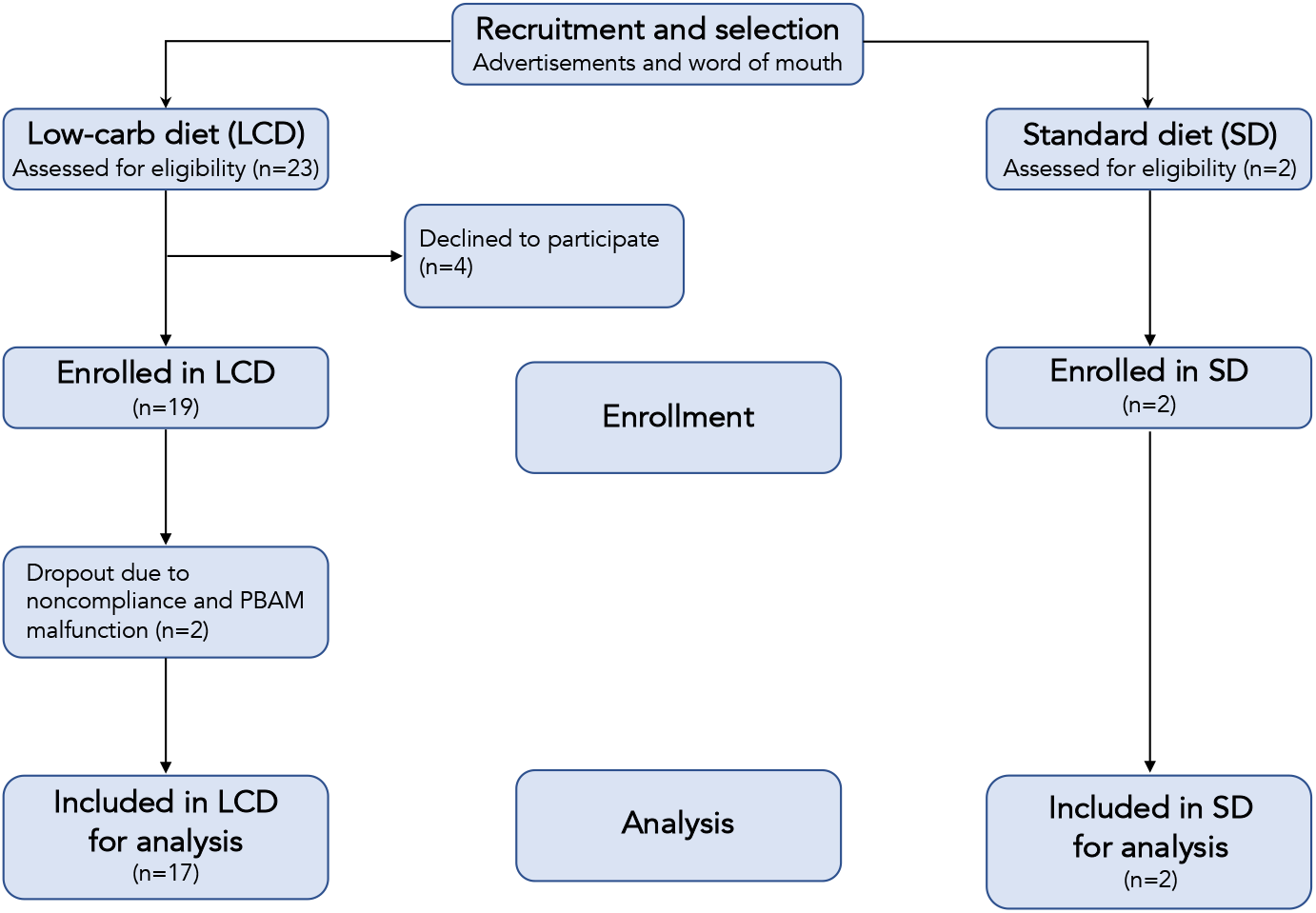
Flow chart of participants in each stage of the study.

### Procedure

All subjects were provided with a PBAM device (BIOSENSE, Readout, Inc.) and a blood ketone meter (Precision Xtra, Abbott Laboratories, Inc.) with test strips and supplies. BIOSENSE is an FDA registered Class I medical device. The Precision Xtra is a commercially available FDA cleared device, and the lancets and alcohol swabs are commercially available medical supplies. Verbal and written study instructions and in-person training on the use of both devices was provided during an initial visit. Subjects were instructed to measure their ketones five times per day every day for two weeks at the following times:

1. First thing in the morning (5 - 9am)
2. Right before lunch (11am - 12pm)
3. Approximately 2 hours after lunch (2 - 4pm)
4. In the evening before dinner (5 - 7pm)
5. In the evening after dinner (8 - 10pm)

At each of the scheduled times, subjects were instructed to take back-to-back breath and blood ketone measurements. Additional breath ketone measurements at other time points during the day were permitted at the subject’s discretion. Subjects were provided with an online form to manually enter their blood and breath ketone values after each measurement session. Because subjects were permitted to select their start date within a 2 week window, the duration of the data collection period was 4 weeks.

Subject were selected according to the diet they were following before the study period, and, therefore, they were not provided with formal dietary guidance. Subjects in the ketogenic diet arm were encouraged to maintain ketone levels at or above 0.5 millimolar (mM) blood BHB, while subjects in the standard mixed cohort were instructed not to adjust their diet based on their ketone readings. This instruction was to prevent behavior change due to non-ketogenic subjects becoming motivated to increase their ketone levels.

### Ethics

This study was approved by the Western Institutional Review Board (Study Number: 1265848) and registered with clinicaltrials.gov (NCT04130724).

Only subjects who had the capacity to provide informed consent were enrolled. The objectives of the study, all experimental procedures, all of the requirements for participation, and any possible discomforts, risks and benefits of participation were clearly explained in writing and orally, in lay terms, to each subject. After all questions were answered, and the subjects were informed orally and in writing that they were free to withdraw from the study at any time with no bias or prejudice, written informed consent was obtained.

### Statistical Analyses

A single daily ketone measurement was compared to multiple daily measurements by calculating the coefficient of variation (CV) for each daily group of measurements for each subject. The magnitude of CV represented the amount of variability in ketone levels during the course of a single day and, by extension, the utility of relying on a single daily ketone measurement to characterize ketosis levels.

The relationship between BrAce and blood BHB was evaluated using several methods. First, a linear ordinary least squares regression analysis was performed to determine the point-by-point correlation between BrAce and blood BHB. A one-sided t-statistic was used to test the null hypothesis that the slope of the regression line was zero. The p-value cutoff for significance was 0.05. Second, a receiver operating characteristic (ROC) analysis was performed to determine the ability of BrAce to predict membership in groups defined by standard blood BHB cutoffs. Example cutoff points were blood BHB values of 0.3 mM (slightly elevated ketones), 0.5 mM (onset of nutritional ketosis), 1.0 and 1.5 mM (higher levels of nutritional ketosis). Sensitivity and specificity of the BrAce test were computed for each blood BHB cutoff. Third, the daily ketone exposure, represented by the area under the curve (AUC) on a ketone concentration versus time graph, was calculated for blood and breath, and the relationship was characterized via linear ordinary least squares regression. For calculations involving cumulative daily metrics like the coefficient of variation (CV) or daily ketone exposure, only days with 4 or 5 measurements were considered. Statistical analyses were performed using standard Python libraries (e.g. NumPy, Pandas).

## RESULTS

Subjects were adherent to the measurement protocol at a rate of 100%, 93% and 63% for 3, 4 and 5 measurements per day, respectively. Data from 2 of the 21 subjects were discarded because of PBAM device malfunction and improper device use. The results presented in this section represent data from the remaining 19 subjects.

To assess the daily variability of ketone concentration at the group level, coefficients of variation (CVs) were calculated by dividing the standard deviation by the mean for each subject-day. Figure 4 shows the probability distribution functions (PDFs) for the daily BrAce and blood BHB CVs. On average, BrAce and blood BHB fluctuate by 43.8% and 45.9%, respectively. In other words, on any given day, a single measurement differs from the time-weighted average by almost 50%. This relatively large variability demonstrates the utility of taking several ketone measurements per day, using either BrAce or blood BHB, in order to capture the full picture of daily ketone dynamics.

**Figure 4.**
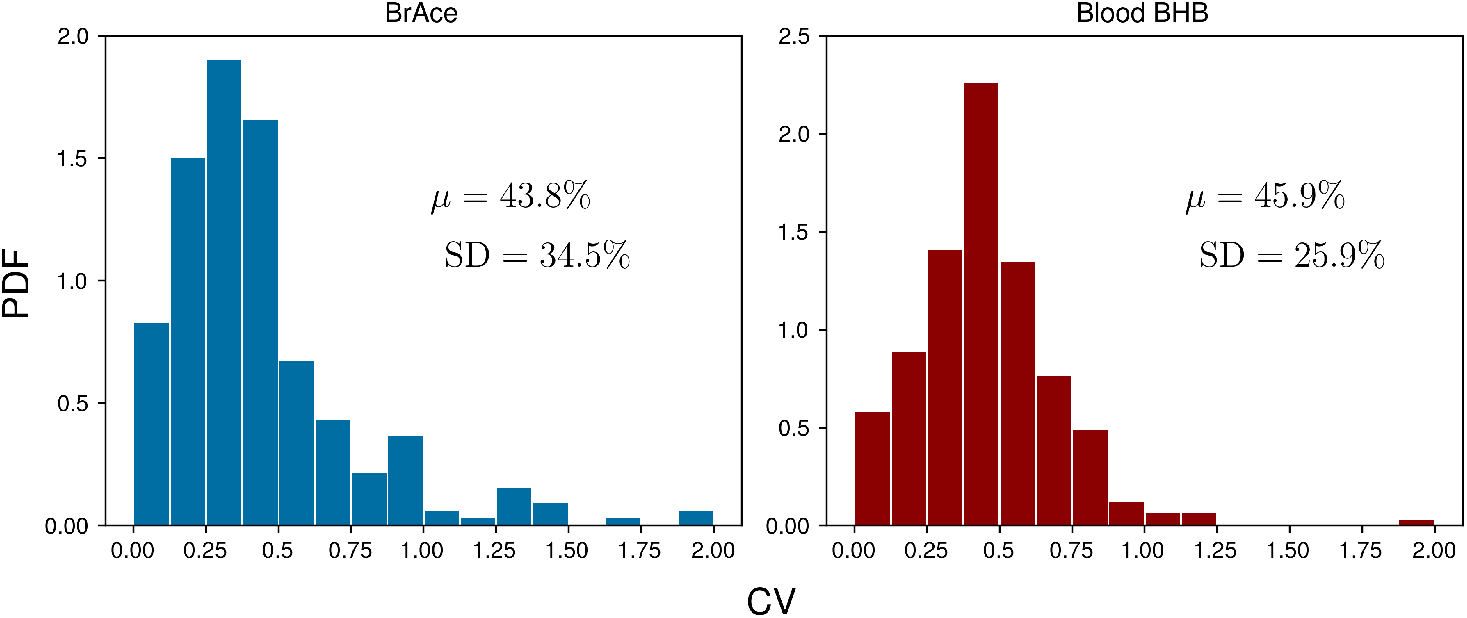
Coefficients of variation (CVs) for each group of daily measurements across all subject-days (n = 248). The mean CV (*µ*) was 43.8% and 45.9% for BrAce and blood BHB, respectively. The similar distributions also imply that BrAce measurements from the PBAM are accurately reflecting the variability in blood BHB.

Linear regression was used to determine the correlation between pairs of coincident BrAce and blood BHB measurements. Figure 5 shows the correlation between coincident BrAce and blood BHB measurements (n=1,214). The strength of this correlation is moderate (*R*^2^ = 0.57) and similar to literature reported values. The grid-like data distribution in Figure 5 is caused by the finite measurement step size of the two ketone meters (1 ACEs for the PBAM and 0.1mM for the blood ketone meter). As a result, there are many overlapping data points in Figure 5, particularly at low ketone levels.

**Figure 5.**
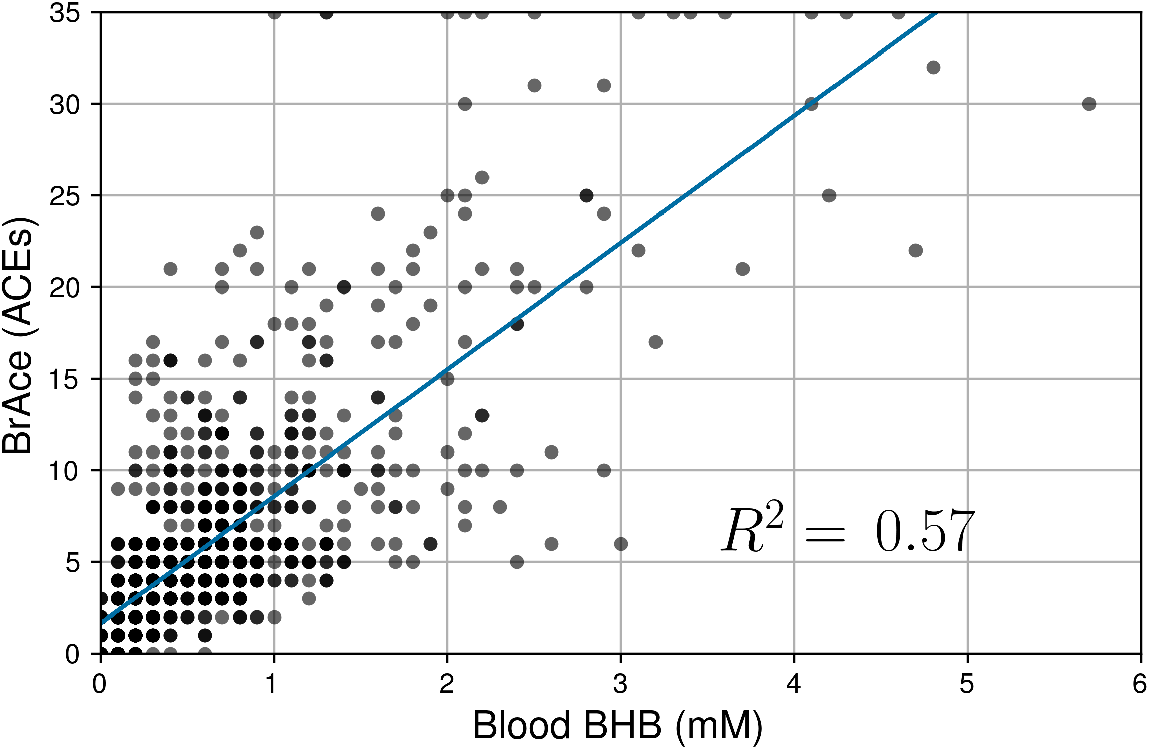
Correlation of coincident BrAce and blood BHB measurements (n=1,214). The gray and black dots represent individual and multiple overlapping data points, respectively. BrAce and blood BHB are linearly correlated with *R*^2^ = 0.57 (P<0.0001). This correlation coefficient is similar to literature reported values whose weighted mean is 0.64.

Next, a receiver operating characteristic (ROC) analysis was performed to determine the diagnostic ability of the PBAM compared to blood BHB, which was treated as the gold standard. For a given BHB threshold *T*, the BrAce measurement was evaluated to either correctly or incorrectly predict membership in the classes BHB ≤ *T* and BHB*> T*. Correct prediction led to a true positive (TP) result while incorrect prediction led to a false positive (FP) result. Performing this analysis for a variety of BrAce levels generated a family of (FP,TP) ordered pairs, which formed an ROC curve (Figure 6).

**Figure 6.**
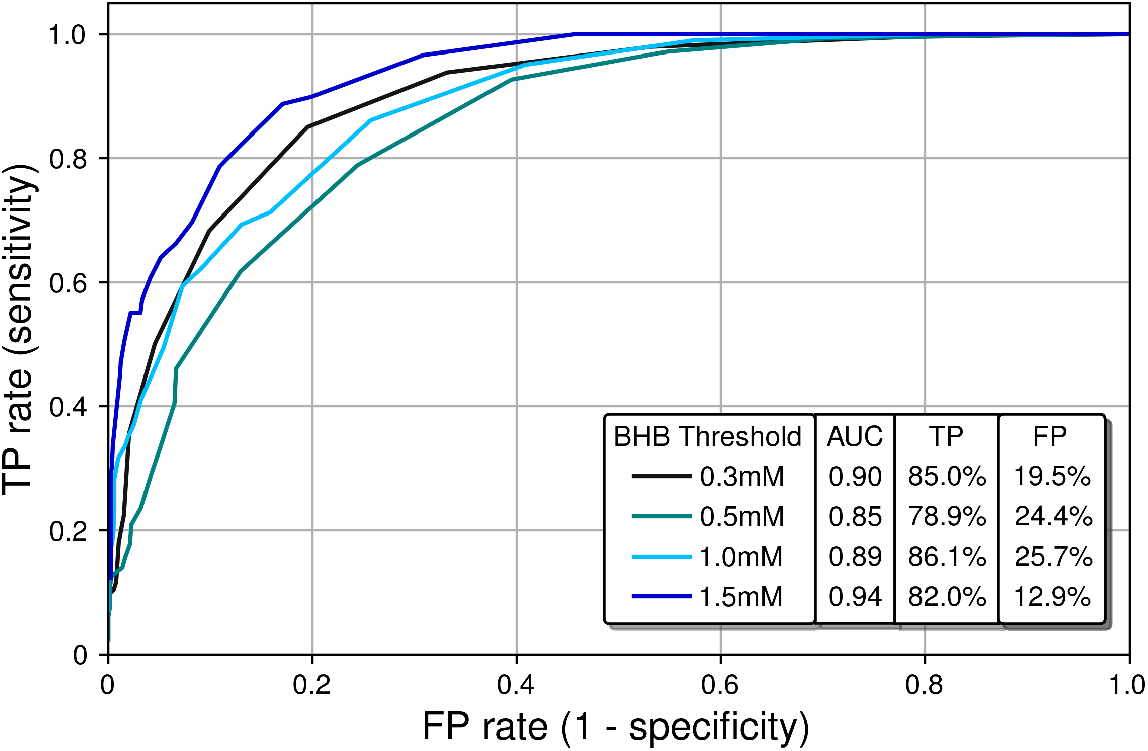
Receiver operating characteristic (ROC) curves characterizing the classification performance of the PBAM BrAce measurement using blood BHB as a standard. AUC calculations indicate that the PBAM measurement is an excellent classifier (AUC *≥* 0.90) of BHB states with 0.3mM and 1.5mM thresholds and a good classifier (AUC *≥* 0.80) for 0.5mM and 1.0mM thresholds.

Ideal classifiers have high TP and low FP rates, which represent data in the upper left quadrant of Figure 6. Classifier quality is typically quantified using the area under the curve (AUC), with AUCs of 1 and 0.5 representing a perfect and a random classifier, respectively. The AUCs in Figure 6 are in the range 0.85-0.94, which demonstrates that the PBAM measurement is either a good or an excellent classifier of blood BHB states depending upon the specific BHB threshold.

Finally, the daily ketone exposure (DKE) for BrAce and blood BHB was calculated and compared. The concept of DKE is borrowed from pharmacology, where the total drug exposure is defined as the area under the curve (AUC) on a concentration versus time graph (Saha, 2018). Analogously, the DKE is calculated from the AUC on a ketone concentration versus time graph. While a single ketone measurement represents the instantaneous concentration of ketones in the body, DKE represents an individual’s cumulative exposure to ketones during a single day.

Figure 7 shows the correlation between DKEs for BrAce and blood BHB (n=248). Breath and blood DKEs were highly correlated with *R*^2^ = 0.83, which implies that PBAM and blood BHB measurements provide similar indications of overall ketone exposure if multiple measurements are taken each day.

**Figure 7.**
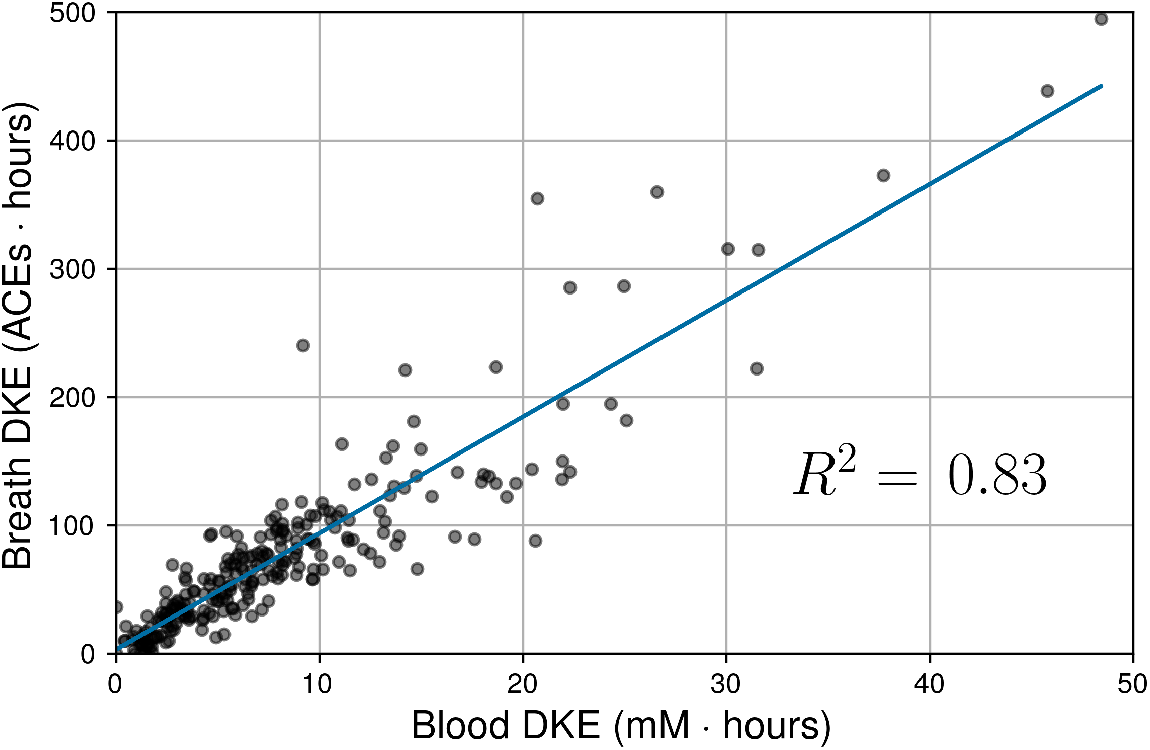
Correlation between daily ketone exposures (DKEs) as measured by BrAce and blood BHB. Each data point represents one subject-day during the trial. The gray and black dots represent individual and multiple overlapping data points, respectively. Blood and breath DKEs were highly correlated

## DISCUSSION

The relatively large variability in daily ketone levels (Figure 4) could be caused by a variety of factors. First, ketone levels are strongly affected by dietary macronutrient composition, with a lower ratio of carbohydrate to fat tending to cause an increase in both BrAce and blood BHB. In addition, ketone levels increase during periods of caloric restriction (Kundu et al., 1993) or fasting (Freund, 1965). Conversely, it has been shown that for subjects on a calorie restricted diet, eating a high carbohydrate snack causes a precipitous drop in BrAce 1-3 hours after the snack is consumed (Kundu, 1990). Although our study did not include dietary tracking, it is likely that daily ketone variability was caused in part by the specific macronutrient content of each individual meal.

Second, vigorous or prolonged exercise can cause an increase in ketone levels in the hours following exercise (Yamai et al., 2009). The rapid depletion of muscle glycogen during exercise and the resulting upregulation of ketogenesis to meet ongoing energy demands can lead to a two-fold increase in ketone levels over the period of several hours (King et al., 2009). Several subjects in the ketogenic/low-carb arm of our study were long distance endurance athletes who reported this pattern when comparing their exercise and ketone logs. In the majority of subject-days, a single daily ketone measurement could not capture this behavior.

Finally, hormones play a role in regulating ketone production, particularly insulin, glucagon and cortisol, which affect the release of FFA from adipose tissue (Alberti et al., 1978). Anecdotally, trial participants frequently observed a drop in BrAce levels upon waking, which may be attributable to increased cortisol levels in the morning and the attendant increase in blood glucose.

The point-by-point correlation between BrAce and blood BHB is moderate (*R*^2^ ∼ 0.6) both in this study and in previously published studies. This result serves to validate the measurement performance of the PBAM as it produces BrAce/BHB correlation results that are comparable with the laboratory mass spectrometer based tools used in the literature. Furthermore, ROC analysis indicates that BrAce measurements from the PBAM can be used to predict whether blood BHB is above or below a certain threshold (Figure 6). The performance of the PBAM as a blood BHB classifier is either good or excellent depending upon the BHB threshold value.

There are multiple factors that may impact the comparison of BrAce and blood BHB, both in the point-by-point correlation and in the ROC analysis. First, the gold standard blood BHB measurement is a laboratory plasma, rather than a point-of-care (POC), test. In fact, a recent study has shown that ketone measurements from POC blood meters often differ from laboratory plasma BHB tests by 0.2mM (Norgren et al., 2020). This same study also demonstrated that results from the POC blood meter were dependent upon whether the sample was derived from capillary or venous blood, with venous blood results matching laboratory plasma tests more closely. These factors may have influenced our study results, which relied on a POC capillary blood meter to gather frequent measurements.

The second factor affecting the BrAce/BHB comparison is the indirect physiological relationship between BrAce and blood BHB. BrAce is not a direct proxy for BHB. Rather, since BrAce is produced directly from AcAc via spontaneous (i.e. unregulated) decarboxylation, the correlation between BrAce and AcAc is higher than that of BrAce and BHB (Musa-Veloso et al., 2002; Rooth and Carlström, 1970). AcAc and BHB, on the other hand, are related through an enzymatically controlled and reversible redox reaction with their relative ratio determined by the mitochondrial redox potential (i.e. NADH/NAD^+^). Since the redox state of the mitochondria is influenced by the energy demands of the body, so too is the instantaneous ratio of AcAc to BHB, and by extension BrAce to BHB. Furthermore, there is evidence that the ratio of AcAc/BHB depends on the overall depth of ketosis (Laffel, 1999). Therefore, there is not always a predictable one-to-one relationship between changes in BrAce and changes in BHB.

The indirect physiological relationship between BrAce and blood BHB also gives rise to interesting temporal dynamics, which in turn affect the correlation and ROC analysis. While BrAce and blood BHB typically changed at similar rates, the concentration extrema were often offset in time. Figure 8 shows example ketone traces from two different subject-days, which demonstrate peak concentrations of blood BHB occurring approximately 4 hours prior to peak concentrations of BrAce. This behavior was evident in 73% of the trial subjects with temporal offsets ranging from 1-5 hours. The remaining subjects showed primarily coincident changes in BrAce and BHB. This temporal offset causes a decrease in the point-by-point correlation coefficient and in the predictive power of the PBAM classifier. Although a time shifted analysis is possible, the variability in the time lag requires that a characteristic time shift be calculated for each subject-day. Because each subject-day contains a small number of data points (4-5), the accuracy and practical utility of such an analysis is limited.

**Figure 8.**
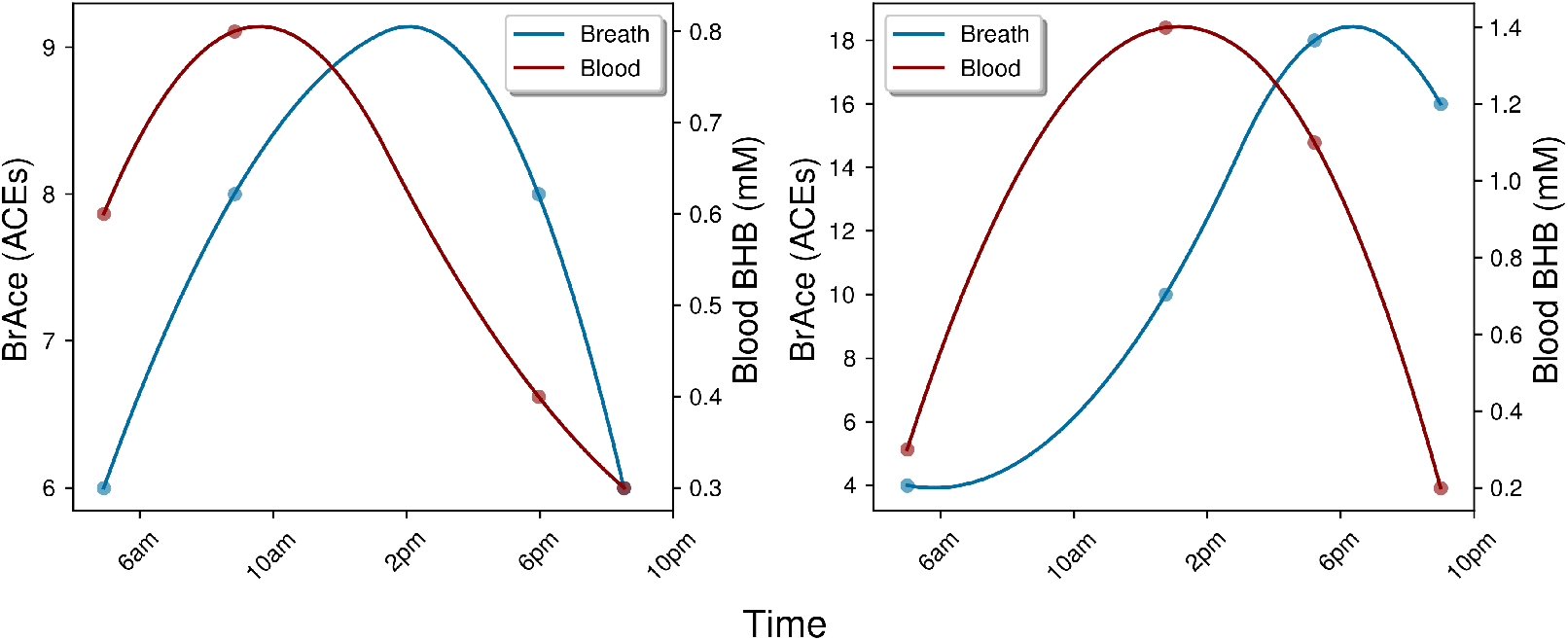
Examples of the temporal lag between blood BHB and BrAce. Both examples demonstrate a lag of approximately 4 hours between peak concentrations of blood BHB and BrAce. This time lag effectively decreases the point-to-point correlation coefficient.

The physiological and biochemical causes of this temporal lag between BrAce and blood BHB are not well understood. However, the processes pictured in Figure 1 can inform a hypothesis. If AcAc is converted to BHB before it has a chance to degrade into acetone, BHB would rise prior to BrAce. As AcAc production continues, either the redox potential and/or the supply of converting enzyme may drop, which would limit additional conversion of AcAc to BHB. The concentration of AcAc would then rise with a portion degrading into acetone and causing an increase in BrAce. In this case the time lag between BHB and BrAce may be an indication of the availability of converting enzyme or the redox potential of the mitochondria.

Point-by-point comparisons of blood and breath are strongly affected by the dynamics of ketone interconversion described above. By contrast, the daily ketone exposure (DKE) is a cumulative daily metric that is less sensitive to such temporal offsets. The high correlation seen in Figure 7 suggests that while instantaneous changes in BrAce and BHB may not always occur simultaneously, the cumulative daily exposure to acetone can be used to predict the daily exposure to BHB and vice versa. Note that while the DKE comparison improves the correlation suppression due to temporal effects, the variable ratio of AcAc/BHB and the limitations of the POC blood test will ensure that the DKE correlation *R*^2^ is less than 1.

## CONCLUSION

These results demonstrate that the PBAM can be used to accurately and noninvasively determine ketosis levels in individuals. The relatively high variability in both BHB and BrAce suggests that single daily measurements are often not sufficient to fully characterize daily ketone exposure. The single time measure correlation between blood BHB and BrAce is moderate and similar to literature values. Furthermore, the PBAM can be used as a diagnostic tool to accurately classify blood BHB states. The PBAM can also be used as a tool to explore the time dynamics that govern changes in BrAce and blood BHB. Those dynamics can be understood by considering the time scales of the interconversion processes that occur between the three ketone bodies. Finally, the daily exposures to BrAce and blood BHB are highly correlated, indicating that the cumulative ketone dose as measured by acetone or BHB are similar.

## Data Availability

Data referred to in the manuscript is available as supplementary file, raw_data_NCT04130724.txt

